# The Mucosal Cytokine Landscape of Acute Gonorrhea Using a Controlled Human Infection Model

**DOI:** 10.64898/2026.02.22.26346846

**Authors:** Michael P. Motley, Marcia M. Hobbs, Andreea Waltmann, Andrew N. Macintyre, Joseph A. Duncan

## Abstract

The host response to *Neisseria gonorrhoeae* is variable, and understanding its systemic and local components is critical to understanding anti-gonococcal immunity for vaccine development. We used a controlled human infection model of male gonococcal urethritis in naïve volunteers in combination with multiplex cytokine analyte analysis of blood and urine specimens taken before infection, at the time of acute symptoms, and after curative treatment of *N. gonorrhoeae* to study responses to early infection. (This study utilized data and specimens from all 11 participants assigned to control arms of two previous randomized clinical trials). All 11 participants developed urethritis between 2 and 5 days post inoculation with *N. gonorrhoeae* strain FA1090, with a majority having visible discharge by day 3. In urine, we found increases in IL-1RA, G-CSF, and chemokines CXCL10, CCL4, CCL11, GROα/β/γ, and IL-8/CXCL8, with IL-1RA and CCL4 showing direct correlation with the degree of pyuria at the time of infection. Contrary to a prior study using the human challenge model and *N. gonorrhoeae* strain MS11mkC, we did not see similar increases in urine IL-6, TNF-α, or IL-1β, although differences in IL-6, TNF-α were observed in participants with later development of infection. Additionally, plasma cytokine levels were unchanged in this cohort over the course of their infection, suggesting these infections were confined to the urethra. We propose that differences in strain virulence or the threshold to define a clinical case may be responsible for this discrepancy, meriting further study and continued use of non-invasive inflammatory markers to study local effects in addition to systemic effects of gonococcal infection.

**Author Summary:** Gonorrhea, caused by the bacterium *Neisseria gonorrhoeae*, remains a global public health concern, yet repeated infections are common and no vaccine is available. A key challenge for vaccine development is limited understanding of how the human immune system responds during early infection, when bacteria are confined to the urethra, vagina, or other mucosal sites. To address this gap, we studied immune responses in a controlled human infection model in which male volunteers with no prior exposure were experimentally infected with *N. gonorrhoeae* into their urethra. Immune signaling molecules were measured in urine and blood samples collected before infection, during symptoms, and after antibiotic treatment.

All participants developed urethral inflammation within a few days of infection. We observed marked increases in multiple inflammatory cytokines in urine, some which correlated with the degree of neutrophils in their urine. In contrast, immune markers in the bloodstream remained largely unchanged. These findings suggest that early infection with the *N. gonorrhoeae* strain tested triggers a strong localized immune response without widespread systemic inflammation. Our results highlight the value of urine-based, non-invasive sampling and demonstrate the power of human challenge models for studying early immune responses that have been difficult to characterize in animal systems.

## Introduction

The human pathogen *Neisseria gonorrhoeae* is a significant cause of morbidity and complication to reproductive health worldwide with over 80 million incident infections occurring annually (1). *N. gonorrhoeae* infects the male and female reproductive tracts, as well as the rectum and oropharynx. Untreated infections can cause pregnancy and birth complications as well as increased risk of HIV transmission. The development of resistance to an increasing number of antibiotics amongst *N. gonorrhoeae* isolates has complicated treatment, necessitating new strategies to treat *N. gonorrhoeae* and prevent it spread. Vaccine development is one promising strategy that has been encouraged by the World Health Organization. Such development requires bolstering our limited knowledge of host immunity and factors required to prevent or respond to establishment of *N. gonorrhoeae* infection within host tissues, as previous attempts at anti-gonococcal vaccines have failed (2–4).

The host response to *Neisseria gonorrhoeae* infection in the reproductive tract varies in severity (5–7). Observational data have shown that pain and discharge associated with symptomatic infection are associated with an influx of leukocytes, predominantly neutrophils, into the urethra or the cervix (8). However, early events that precede the development of *N. gonorrhoeae* infection, and factors that predispose development of symptomatic infection, asymptomatic infection, or clearance of *N. gonorrhoeae* without infection are not well studied. In nature, *N. gonorrhoeae* is exclusively a human pathogen. Thus, most studies of immune response to infection with *N. gonorrhoeae* are limited to observational cohorts of patients with active *N. gonorrhoeae* infection and/or historical infection. Mouse models can simulate but not fully recapitulate gonococcal infection, and to date there is not an accepted model of male gonococcal urethritis in mice (9–11). Increasing data have revealed differences in mucosal immunity compared with systemic immune responses that may be crucial to early defense against infection. For example, the human urethra is found to have resident T lymphocyte populations, including CD8 cells, as well as active plasma cells (12, 13). Thus, there is a need to study local human responses to *N. gonorrhoeae* infection.

The controlled human infection model (CHIM) of male gonococcal urethritis has been used by our group for over thirty years investigating mechanisms of *Neisseria gonorrhea* pathogenesis within the human male urethra (14, 15). In this model, healthy male volunteer participants undergo urethral inoculation with *N. gonorrhoeae* followed by daily monitoring for development of clinical urethritis and subsequently treatment for infection once symptoms arise. The model enables prospective sampling before, during, and after infection, permitting temporal resolution when analyzing immune dynamics. Because all individuals in a study are exposed to a specific strain of *N. gonorrhoeae*, biologic variability in host response due to strain variation is controlled. Additionally, volunteers have been screened for history of *N. gonorrhoeae* infection, so all observed responses are the naïve immune response to the pathogen.

While the CHIM used to date has not included invasive sampling, such as the collection of tissues, bodily fluids such as blood, semen, and urine have been collected in studies and used for biologic assays. First-catch urine provides a sampling of the infected mucosal surface that includes bacteria, immune cells, and soluble immunologic mediators like cytokines (16–18). One study using CHIM to study *N. gonorrhoeae* in male volunteers examined changes in the concentrations of six cytokines in urine and plasma, as well as the corresponding mRNA levels in circulating PBMC, before infection with *N. gonorrhoeae* strain MS11mkC and at time of symptoms. This study observed elevations in urine Interleukin (IL)-8 (aka CXCL8), IL-6, Interferon (IFN)-γ, and IL-1β in response to infection, with plasma levels of these cytokines also increasing at symptom onset in a subset of participants(7). In this study, we further this previous work characterizing the cytokine responses to gonococcal infection using controlled human infection with a second strain of *N. gonorrhoeae* and examining a more extensive panel of cytokines. Our study demonstrates that gonococcal urethritis can occur without systemic immune responses, and that local chemotactic factors are elevated in response to early infection.

## Results

### Infection dynamics after inoculation with *Neisseria gonorrhoeae* FA1090 demonstrate development of gonococcal urethritis in all study participants

Between 2014 and 2015, and between 2017 and 2018, eleven male volunteers, 18-35 years of age without prior *N. gonorrhoeae* exposure, were inoculated with wild type *N. gonorrhoeae* strain FA1090 under clinical trials NCT04870138 and NCT03840811 using our well-established protocol (Fig. 1A) (19). The participants were monitored daily for symptoms, and first-void urine was collected daily to longitudinally assess the level of bacteriuria (measured as CFU/mL of cultured urine) and degree of pyuria in the urine sediment (measured as cells/mL sediment by microscopy) (19). All participants demonstrated bacteriuria on day 1 post-inoculation, had progressively higher urine bacterial counts on each subsequent day, and all developed clinical gonococcal urethritis, defined as visible urethral discharge observed by the study clinician, by day 5 of the study (Fig. 1B). All study participants were administered curative antibiotic therapy when urethral discharge was first observed by the study clinician. Six participants (55%) developed urethritis within 2 days of inoculation; the average time to urethritis development was 2.6 days. Bacterial burden and urine sediment neutrophil counts at time of urethritis differed by participant (2.0 – 6.4 log_10_ CFU/mL and 5.8 – 7.0 log_10_ PMN/mL sediment, respectively) (Fig. 1C, 1D). Follow up visits within a week of treatment confirmed elimination of *N. gonorrhoeae* with nucleic acid testing and resolution of urethritis on clinical exam for all study participants.

**Fig 1.**
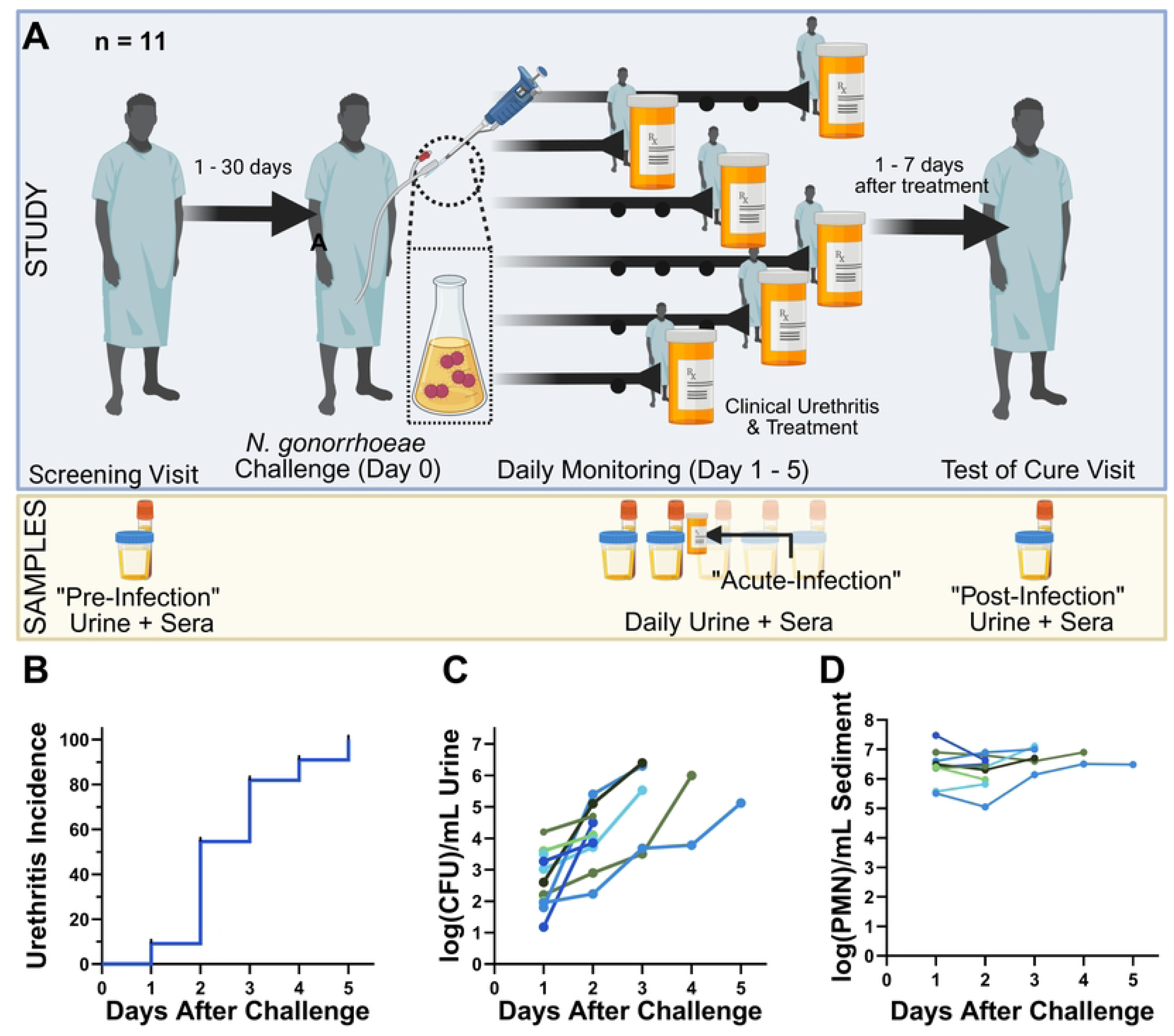
Neisseria gonorrhoeae urethritis CHIM Model and Summary Results. (A) Schematic of the experimental infection model, with annotations of when urine and blood was collected during the experiment; (B) Time to urethritis post-challenge for all 11 participants; (C) Urine bacterial burden and (D) Urine sediment neutrophil counts at each visit post-challenge. Each color represents a different participant. Created in BioRender. Motley, M. (2026) https://BioRender.com/aiujxhc

### Urethral infection with *Neisseria gonorrhoeae* causes increases in urine monocytic and granulocytic chemokines from baseline

We collected plasma and first-void urine samples at each participant’s screening visit (up to thirty days prior to inoculation, hereafter referred to as Pre-Infection); at the visit that each participant demonstrated clinical urethritis (hereafter referred to as Acute-Infection); and at each participant’s test of cure visit (hereafter referred to as Post-Infection). These samples were used to analyze cytokines present at these three time points for each participant. 41 cytokines were measured in urine, and 38 cytokines were measured in plasma. Much of the data in our study fell outside the limits of quantitation; in urine, only 30% of cytokine values fell within the limits of quantitation. For one cytokine (Epidermal Growth Factor), 25 of 33 data points (76%) from urine samples were above quantitation limit defined by our standards and was thus excluded from further analysis. For 7 urine cytokines (IL-10; IL-13, IL-15, IL-1α; IL-2; IL-3; IL-5), all 33 (100%) data points were below the lowest standard measured which we hereafter refer to as the lower limit of quantitation (LLOQ). An additional 18 cytokines (TGF-α; GM-CSF; IFN-α2; IFN-γ, CCL7, IL-12p40; CCL22, IL-12p70; sCD40L; IL-17A, IL-9; IL-1β; IL-4, IL-6, IL-7, CCL3, TNF-α, and TNF-β) had a median concentration below the LLOQ for all 3 time points. Independent data points are shown graphically for these cytokines in **S1 Fig**, but further formal statistical analysis for these analytes was limited. The remaining 15 cytokines underwent hypothesis testing (Figure 2, Table 1). These 15 cytokines consist primarily of growth factors involved in wound healing (e.g. FGF2, PDGF, VEGF) and chemoattractants in pro-inflammatory innate immunity (e.g. G-CSF, CX3CL1, GRO/CXCL1/2/3, IL-8/CXCL8, CXCL10). All values are reported in **S1 Table**.

**Figure 2.**
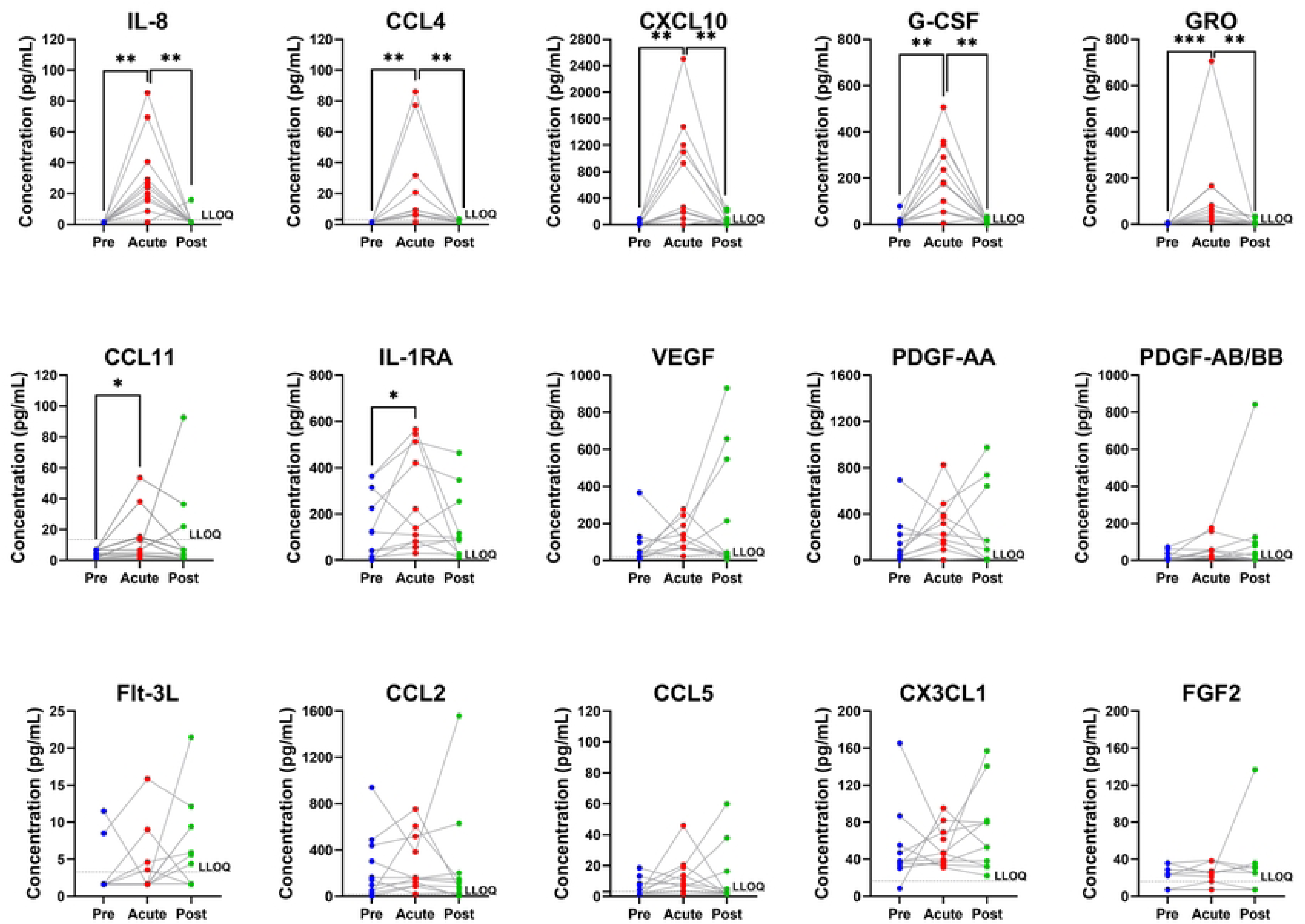
Comparison of urine cytokines at each timepoint. Each dot represents a cytokine measurement from one participant, with lines connecting measurements from the same participant. Values below the lower limit of Quantitation (LLOQ, dotted line) are displayed as one half LLOQ for and unresolved from other non-detects. Comparison bars indicate statistical significance at *p<0.05; **p<0.01 via paired Wilcoxon-signed rank test between Pre- and Acute-Infection cytokines, and between Acute-and Post-Infection cytokines.

**Table 1.** Cytokine Concentrations of those values with the majority of medians above the lower limit of quantitation (LLOQ). Median values below the lower limit of quantitation (LLOQ) are indicated. If both the first and third quartiles are below the limit of detection, no interquartile range (IQR) is given. Statistical testing for each pair of cytokines tested was performed using two-tailed paired Wilcoxon Signed Rank test using Pratt correction for matched, censored values. Censored values were substitute for half the LLOD for statistical purposes. *For Eotaxin, the Lower Limits of Detection for the two assays that were run were so dissimilar that the median was below one of the two limits of detection

|  | Pre Infection |  | Acute Infection |  | Post-Infection |  | p value | p value |
| --- | --- | --- | --- | --- | --- | --- | --- | --- |
| Cytokine | Median | IQR | Median | IQR | Median | IQR | Pre vs Acute | Acute vs Post |
| Fibroblast growth factor 2 (FGF2) | 22.34 | LLOQ- 29.00 | 21.31 | LLOQ- 27.18 | 25.30 | LLOQ- 32.48 | 0.625 | 0.906 |
| Eotaxin* (CCL11) | <LLOQ | LLOQ- 6.80 | <LLOQ | LLOQ- 15.43 | <LLOQ | LLOQ- 21.91 | **0.016** | 0.258 |
| Granulocyte Colony-Stimulating Factor (G-CSF) | 9.53 | 7.30- 18.60 | 181.49 | 55.81- 343.60 | 11.40 | LLOQ- 17.87 | **0.002** | **0.002** |
| FMS-related tyrosine kinase 3 ligand (Flt-3L) | <LLOQ | N/A | <LLOQ | LLOQ- 4.60 | 5.60 | LLOQ- 9.40 | 0.375 | 0.258 |
| Fractalkine (CX3CL1) | 37.60 | 31.37- 55.23 | 46.13 | 36.47- 69.40 | 53.74 | 32.23- 82.00 | 0.700 | 0.492 |
| GROα/$\beta$/$\gamma$ (CXCL1/2/3) | <LLOQ | LLOQ- 3.40 | 53.48 | 18.70- 164.10 | 4.30 | LLOQ- 10.80 | **0.001** | **0.001** |
| Platelet-derived growth factor AA (PDGF-AA) | 50.80 | 19.99- 225.60 | 227.60 | 145.05- 391.40 | 94.40 | 7.23- 642.37 | 0.148 | 0.638 |
| Platelet-derived growth factor AB/BB (PDGF-AB/BB) | <LLOQ | LLOQ- 38.90 | 26.79 | LLOQ- 57.24 | 26.79 | LLOQ- 95.11 | 0.074 | 0.898 |
| Interleukin 1 Receptor Antagonist (IL-1RA) | 122.30 | 16.00- 315.16 | 139.64 | 79.40- 512.29 | 93.80 | 28.30- 254.96 | **0.024** | 0.102 |
| Interleukin 8 (IL-8/CXCL8) | <LLOQ | N/A | 24.00 | 15.50- 40.53 | <LLOQ | N/A | **0.002** | **0.005** |
| Interferon-gamma-induced protein 10 (IP-10/CXCL10) | 9.47 | LLOQ- 24.90 | 265.50 | 185.90- 1200.40 | 19.90 | LLOQ- 90.47 | **0.002** | **0.002** |
| Monocyte Chemoattractant Protein 1 (MCP-1/CCL2) | 144.31 | 25.40- 439.18 | 154.00 | 95.40- 518.24 | 118.10 | 32.29- 201.80 | 0.577 | 0.520 |
| Macrophage Inflammatory Protein 1 beta (MIP-1b/CCL4) | <LLOQ | N/A | 9.06 | 5.90- 31.75 | <LLOQ | N/A | **0.004** | **0.004** |
| Regulated on Activation Normal T-cell Expressed and Secreted (RANTES/CCL5) | <LLOQ | LLOQ- 8.40 | 8.40 | 3.60- 18.94 | <LLOQ | LLOQ- 16.40 | 0.141 | 0.606 |
| Vascular Endothelial Growth Factor (VEGF) | 46.90 | 18.50- 97.58 | 112.24 | 68.60- 189.33 | 42.20 | 25.70- 547.23 | 0.175 | 0.700 |

Amongst these 15 cytokines, 7 demonstrated significant elevations between Pre-Infection baseline levels and Acute-Infection levels using an alpha of 0.05 (Table 1) (Fig 2). All but one of these cytokines (IL-1RA) are associated with chemoattraction and survival of immune mediators, particularly monocytes (CXCL10, CCL4), granulocytes (CCL11, GRO, G-CSF, IL-8/CXCL8), T cells, and NK cells (CXCL10). Additionally, 5 of these 7 cytokines (CCL4, CXCL10, IL-8/CXCL8, GRO, G-CSF) had Post-Infection levels that showed significant returns to baseline after *N. gonorrhoeae* cure at an uncorrected alpha of 0.05.

### Pre-infection urine cytokine content did not predict time to urethritis presentation, but elevated urine CCL4, IL-1RA, TNF-α, and IL-6 at acute infection were associated with delayed presentation

Volunteers developed urethritis on different days following gonococcal challenge as expected based on data from prior challenge experiments. We subsequently queried if development of urethritis early or later after challenge correlated with differences in urine cytokine concentrations prior to infection. We divided participants into those that experienced urethritis early (Day 1 – 2 after inoculation) versus participants who experienced infection later during follow up (Day 3 – 5) and compared each group’s baseline Pre-Infection levels of cytokines (to evaluate if any “predisposing” signatures were present). We recognized that some of the 18 cytokines with median values below the LLOQ may be disproportionately present in one of these groups, and so these 18 analytes were included in this analysis despite the limited data stratification. No significant differences were observed in baseline Pre-Infection cytokine levels between early and late groups (data not shown), failing to support the hypothesis that baseline cytokines may predict better host defense and therefore delayed urethritis.

We then repeated this analysis using the urine cytokine levels measured during acute infection and found that urine levels of CCL4, IL-1RA, TNF-α and IL-6 at acute infection were all significantly higher in those developing urethritis later when compared with those who developed urethritis early (Mann-Whitney U Test, p=0.030, p=0.030, p=0.028, and p=0.015, respectively) (**Fig. 3**). Unfortunately, we did not assess urine cytokines after inoculation but prior to symptom onset, and so we could not perform a longitudinal analysis with matched timepoints between the early and late onsets. Other comparisons are provided in **S2 Fig**.

**Fig. 3.**
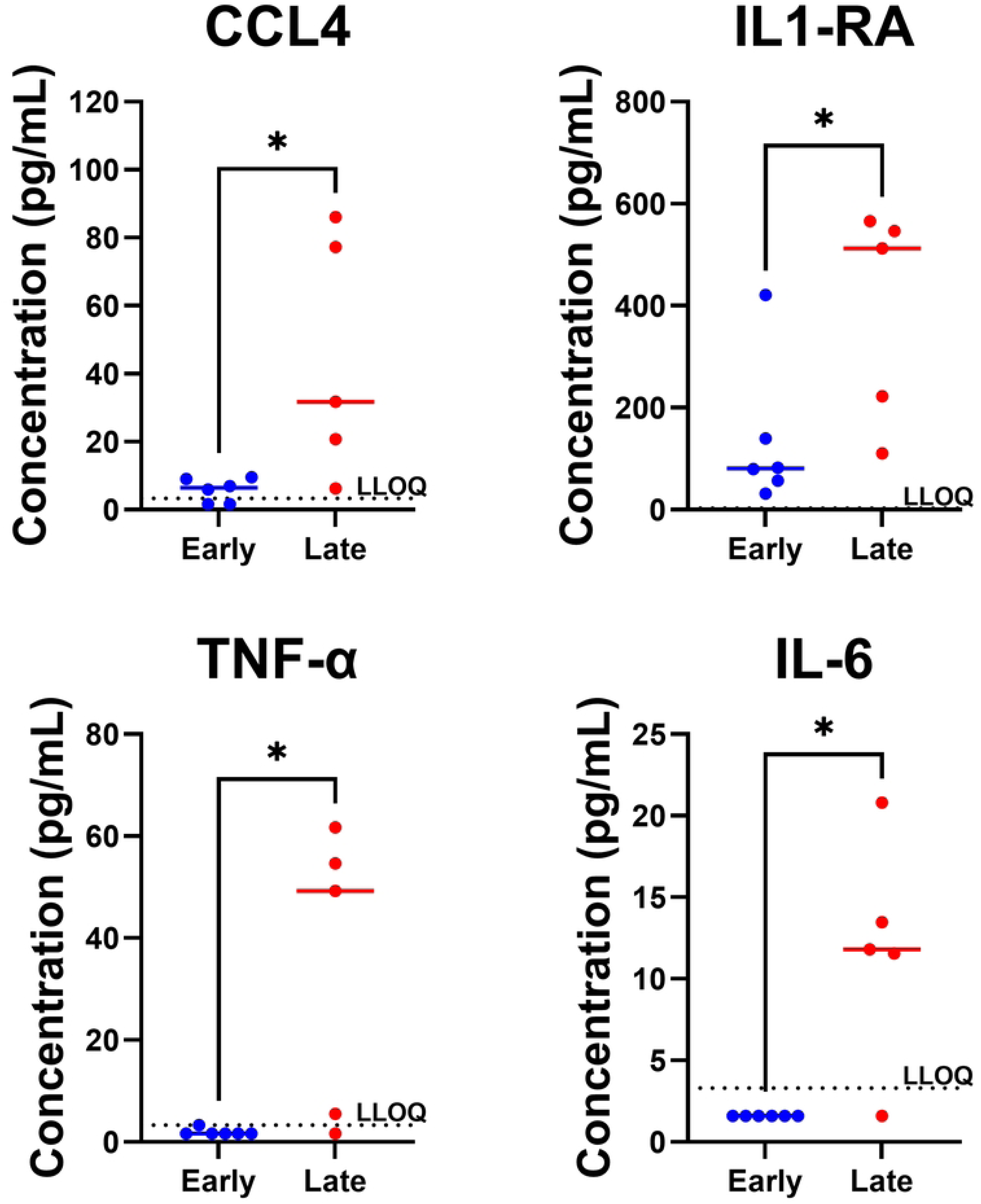
Comparison in Acute Infection Urine Cytokine Levels in Early and Late Presenters. Each dot represents a cytokine measurement from one participant. Values below the lower limit of Quantitation (LLOQ, dotted line) are displayed as one half LLOQ. Comparison bars indicate statistical significance at *p<0.05; via Mann-Whitney U Test.

### Both pro- and anti-inflammatory cytokines in urine are associated with infection severity

The quantity of neutrophils and bacteria present within urine of patients with gonococcal urethritis have both been previously correlated with severity of infection (8, 20). To assess whether urine cytokine levels also reflect infection severity, we performed correlation testing to evaluate the relationships between urine bacterial burden at time of clinical urethritis (expressed as log CFU/mL), neutrophil counts in urine sediment (expressed as log PMN/mL), and each cytokine (expressed as log pg/mL) at the same timepoint. Bacterial burden explained a moderate fraction of variation in urine sediment neutrophil counts (R² = 0.36), although this association did not meet the prespecified significance threshold (p = 0.052, Figure 4). We did find correlation between urine sediment neutrophil counts and PDGF-AB/BB (R^2^ = 0.384, p = 0.042), IL-1RA (R^2^ = 0.581, p = 0.006), and CCL4/MIP-1β (R^2^ = 0.437, p = 0.027), **Fig. 4**). Other tested correlations are in **S3 Fig**.

**Fig. 4.**
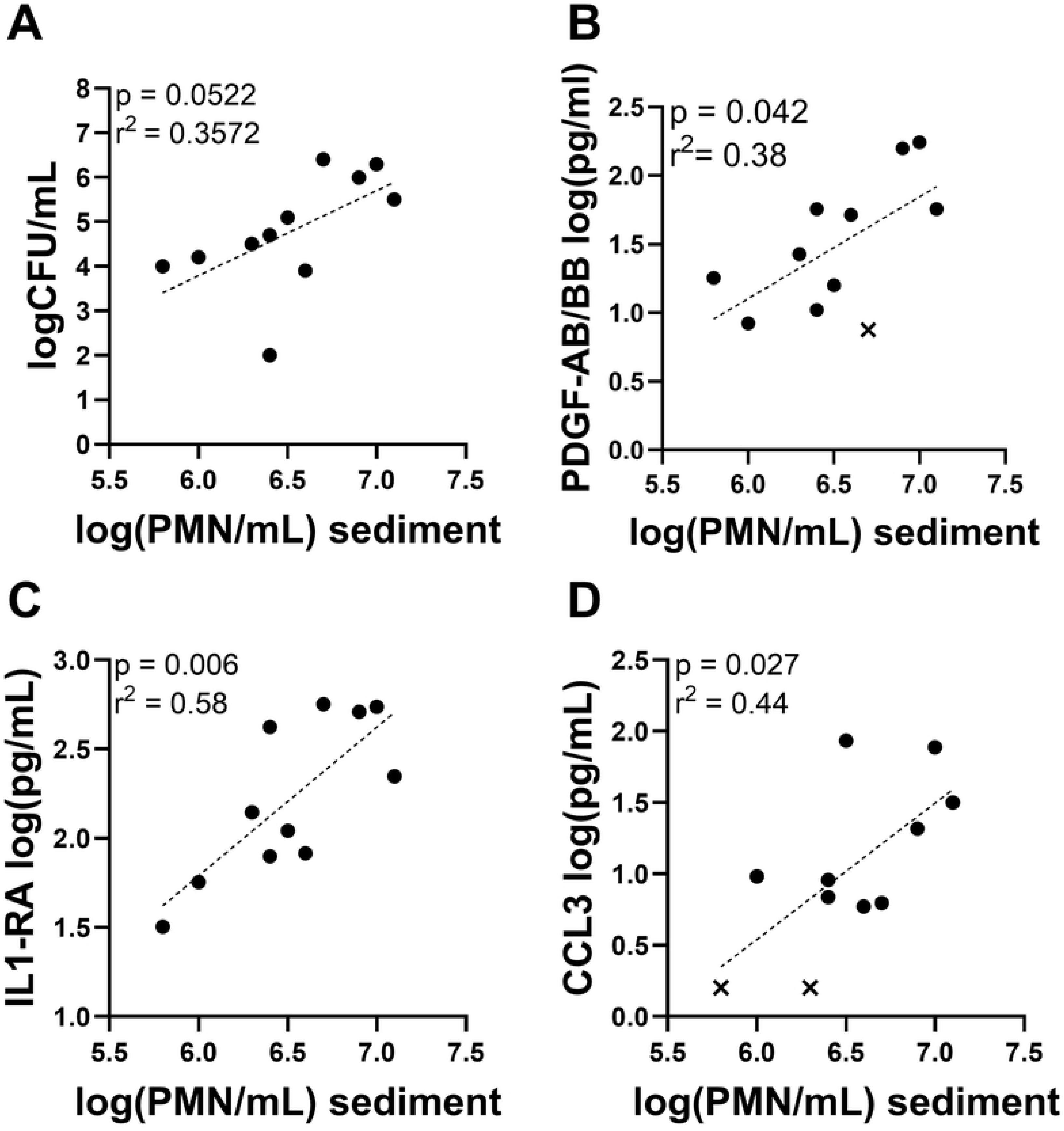
Linear correlation between urine measurements of all participants. (A) Urine sediment neutrophil counts vs Bacterial Burden; (B) Urine sediment neutrophil counts vs Urine PDGF-AB/BB at the acute infection timepoint. (C) Urine sediment neutrophil counts vs Urine IL1-RA; (D) Urine sediment neutrophil counts vs urine CCL3 at the acute infection timepoint. All values are log transformed. Symbols represent individual patients. Individual values below the lower limit of Quantitation are represented with an “x” rather than a dot. Also displayed are results of linear correlation testing, both the p value of significance as well as r2 or goodness of fit. Slopes were not constrained to include the origin.

### Circulating cytokines did not change significantly during acute urethral infection

Compared to urine, cytokines in the plasma were more readily quantified, with more than 50% of the values for 28 of the 38 cytokines tested falling within the range of quantitation for the assay. However, across the course of the infection the mean plasma concentrations of these 28 analytes did not change significantly (Supplemental Figure 4). Moreover, within-individual cytokine profiles remained largely stable over time, with minimal variation observed before, at the onset of clinically apparent infection, or after infection. Such findings suggest there was not a robust acute systemic cytokine response to the presence of early infection in these volunteers. Stratifying participants into early and late presenter groups as above did uncover that late presenters did have higher levels of plasma IFN-α2 (p=0.0390) at Pre-Infection baseline compared to early presenters.

## Discussion

In this study, we describe the cytokine response to experimental urethral *N. gonorrhoeae* infection by wild-type strain FA1090 in naïve human male volunteers). This work builds on prior work that had examined urinary and plasma levels of a handful of cytokines (IL-8, IL-6, TNF-α, IL-1α, IL-1β, GM-CSF) in response to experimental human infection with *N. gonorrhoeae* strain MS11mkC(7).

In general, we found that development of urethritis caused by *N. gonorrhoeae* strain FA1090 was associated with significant elevations in urine levels of CXCL10, CCL4, CCL11, GRO, IL-8/CXCL8, G-CSF, and IL1-RA. Five of these seven inflammatory factors are chemokines associated with inflammatory cell invasion. These cytokines are often associated with urethritis caused by other pathogens and have also been observed in cervical tissues in response to other sexually transmitted infections, such as *Chlamydia trachomatis* (21, 22). Increased CXCL10 and IL-8 expression have also been demonstrated in cervical epithelium infection with *N. gonorrhoeae in vitro* (*23*). GRO and CCL2 have also been found to be increased in early responses to *in-vitro N. gonorrhoeae* exposure (24). Our data are consistent with these findings and suggest that responses to gonococcal urethritis are consistent with general innate immune responses to other bacterial pathogens that cause urethritis.

Additionally, we found only limited detection of TNF-α, IL-6, IFN-γ, or IL-1β in urine even at time of acute infection, although urine TNF-α and IL-6 were found to be elevated in a subset of patients who developed urethritis later. Our study contrasts with a prior reported study by Ramsey *et al.*, which performed a similar controlled human infection model (CHIM) utilizing *Neisseria gonorrhoeae* strain MS11mkC and demonstrated measurable and significant increases in TNF-α, IL-6, and IL-1β on the day of clinical urethritis (7).

We propose several hypotheses regarding this discrepancy. Firstly, FA1090 and MS11mkC are known to differ in infectivity, with the latter possessing a genetic island encoding a type 4 secretion system that lowers the necessary infective inoculum (ID50). As a result, the participants in the Ramsey *et al.* study were inoculated with 50-500 fold less CFU than participants in this study, which may trigger a different immune response. Secondly, the ELISAs used by Ramsey *et al.* reportedly had lower limits of detection than the LLOQ of the Luminex assays used here, which may explain why Ramsey et al. reported detectable urine IL-1β, IL-6, TNF-α and IL-8 in all participants at time of acute infection, while IL-8 was the only cytokine we consistently detected in urine at time of acute infection. The absolute concentrations of the urine cytokines we measured were also generally lower than those reported by Ramsey and colleagues. However, neither sets of assays included independent standards to assess accuracy, and incongruities in absolute results between immunoassay platforms and vendors have been widely documented by our group and others(25–28). Thirdly, the Ramsey study performed more frequent observations after inoculation with patients instructed to urinate and consume fluids regularly, which may limit accumulation of urethral discharge that defines the clinical outcome. In contrast, our study assessed patients only daily, with first-void urine only as the study specimen. It’s possible our approach allowed for the overnight buildup of discharge, prompting earlier diagnosis and treatment before further inflammation could occur. Consistent with this hypothesis, the time to development of symptoms and treatment in the Ramsey study was 62-146 hours post inoculation, while time to symptoms driving treatment for our study was 24-120 hours. Of note, Ramsey *et al* observed that urine IL-8 levels increased within hours of infection – faster than either TNF-α or IL-1β – and so our “acute infection” samples may have been representative of an earlier phase of the immune response.

The last hypotheses are supported by our findings that participants who developed later urethritis, between 72 and 120 hours post inoculation, demonstrated elevated urine IL-6 and TNF-α compared to those individuals who developed early urethritis 24-48 hours after inoculation. Thus, these canonical inflammatory cytokines may be part of the natural pathogenesis of infection but may arise later in the course of infection. Additionally, we found 10-fold less *N. gonorrhoeae* CFU recovered in urinary sediment from participants in our study compared with CFU recovered from those infected with MS11mkC in the Ramsey study. Given the weak correlation between CFU and pyuria and cytokine levels in our study, we propose that the higher counts of bacterial recovered at the time of infection in Ramsey *et. al*. may have also improved detection of IL-6, TNF-α, and IL-1β

The origin of the urine cytokines we detected is unclear and difficult to resolve because of the intertwined, dynamic nature of cytokine production and mucosal tract immune responses. The cytokines could be secreted from the urethral tract of infected men. Alternatively, they could be derived from inflammatory cells or epithelial cells that are shed into the urine as urethral exudate is bathed in initial urine flow (29). A study of inflammatory monocytes showed that stimulation with LPS and peptidoglycan was associated with high expression of chemokine genes including IL-8 and MIP-1β, in both cases more so than expression levels of IL-1β or IL-6 (30). Furthermore, direct exposure of human monocytes to *N. gonorrhoeae* also causes early, inoculum dose-independent stimulation of IL-8, IL-6, GRO and CCL2 followed by later inoculum dose-dependent expression of IL1β, TNF-α and IL-10 (24).

Our data suggest association between pyuria and bacterial load, although these findings were not sufficiently powered to be statistically significant. Additionally, we demonstrate correlations between degree of pyuria and the amounts of urine CCL4, PDGF-AB/BB and IL-1RA. CCL4, which promotes chemotaxis of macrophages and natural killer cells, is potentially a product of the neutrophils initially recruited by locally elevated IL-8. Macrophages have been associated with gonococcal infections, but have been shown to harbor *N. gonorrhoeae* without killing it, which may be due to gonococcal virulence factors. In addition to its association with pyuria, we found that those participants who presented later with urethritis had higher urine CCL4 at the acute infection timepoint than those who presented earlier. It is unclear whether increased CCL4 presence, or the increased presence of IL-1RA, TNF-α, and IL-6 also observed in later presentations, are associated with a more robust host response slowing infection progression, or as discussed above, simply indicative of natural progression of the inflammatory response to *Neisseria gonorrhoeae* if left untreated. PDGF-AB/BB, which is produced by multiple cell types and is a neutrophil attractant may be an indicator of gonococcal-induced damage, and the correlation is intuitive. Interpreting the correlation between pyuria and IL-1RA is more complex. IL-1RA is an acute phase protein that is produced both by myeloid line cells such as monocytes, macrophages, and neutrophils (31). It blocks IL-1α and IL-1β signaling through the IL-1Receptor and so constrains immune activation and prevents collateral damage. However, this can be counterproductive as elevated IL-1RA is also associated with worse severity of pneumonia and other infections (32, 33). Here, it is unclear whether IL-1RA is a response to increased inflammation and a product of the pyuria, or if expression of IL-1RA by other factors (such as through manipulation of host responses by *N. gonorrhoeae*) causes worse severity of infection that is indicated by the pyuria. Further studies of the interaction between *N. gonorrhoeae* with urothelium and IL-1RA are merited.

Compared with our urine data, we did not find changes in plasma cytokines, with variable levels across participants that were unchanged at the Acute Infection and Post-Infection timepoints. This differs from data from Ramsey *et al.*, who observed increases in plasma IL-8 in nearly all participants upon acute infection, plus increases in plasma IL-1RA, TNF-α, and IL-1β in a subset of individuals. These differences could similarly be related to differences in virulence between FA1090 and MS11mkC or differences in the observation of participants and diagnosis of clinical urethritis that may have indicated earlier treatment before these systemic responses could occur(14). It has been demonstrated that challenge with MS11mkC can lead to the induction of plasma and urine antibody responses to *N. gonorrhoeae* outer membrane proteins in some participants two weeks after urethritis (28). Unfortunately we did not collect additional follow-ups beyond the test-of-cure visit, and so we could not assess if challenge with FA1090, and the relatively modest systemic response, was sufficient to induce an antibody response.

In summary, we demonstrate that local immune responses to early infection by *Neisseria gonorrhoeae* can be studied by non-invasive sampling of urine, although detection of the full cytokine response may lag behind the first signs of urethritis. We found that urine G-CSF, GRO (CXCL1/2/3), IP-10, MIP-1β rise early, highlighting the role of these chemokines in early local immune responses, whereas other cytokines were exclusively present in urine during later presentations of infection. However, because our Controlled Human Infection Model protocol requires immediate treatment at the first sign of urethritis, we are limited in the ability to assess cytokine changes associated with further progression of urethritis. Additionally, the current protocol prevents evaluation whether differences in cytokine production are associated with natural clearance or resistance to infection. In our study, all patients developed urethritis upon challenge, whereas Ramsey *et al.* identified several individuals who did not sustain infection after urethral challenge with MS11mkC *N. gonorrhoeae*. However, cytokine responses in these participants was not reported. Future studies could potentially extend the period between symptoms and infections, as was performed by Schmidt and colleagues in attempt to induce a protective antibody response (34). Alternatively, a natural history study of asymptomatic *N. gonorrhoeae* infections, or a model using an inoculum that yields a higher percentage of uninfected participants may allow for a more comprehensive query of differences in urine cytokines in respect to protection against infection. Finally, we are currently studying the utility of detecting RNA within the urine of CHIM participants, which may allow for earlier detection of immune activation. Whether there was a transient, weak systemic cytokine response within the few day window between infection and acute symptoms remains to be evaluated.

Studying the difference between local immune responses and systemic immune responses and how these may play a role in natural infection is important in rational vaccine development against *N. gonorrhoeae*. As additional controlled human infection models for *N. gonorrhoeae* emerge, such as an oropharyngeal model, use of non-invasive sampling to evaluate local responses and systemic responses will be important.

## Methods

### Ethics and Regulatory statements

Written informed consent was obtained from each participant prior to conducting any study activities. All experiments were performed under either NCT04870138 (retroactive registration date 2021-04-29 with recruitment dates 2013-03-10 to 2015-04-28) or NCT03840811 (registration date 2019-02-07 with enrollment dates 2017-04-23 to 2019-11-25) according to the guidelines of the U.S. Department of Health and Human Services and with approval by the University of North Carolina at Chapel Hill Institutional Review Board (approved protocol 12–0482) under an investigational new drug (IND) authorization by the U.S. Food and Drug Administration (IND 15123).

### Experimental Human Infections

Experimental human infections were conducted between 2015–2018 in the Clinical and Translational Research Center of the North Carolina Translational and Clinical Sciences Institute at the University of North Carolina at Chapel Hill as part of two randomized controlled trials (NCT04870138, NCT03840811, (35, 36)). The participants in this secondary analysis were members of the control arms, healthy male volunteers 18-35 without history of prior *N. gonorrhoeae* infection (general inclusion and exclusion criteria for our human challenge model are found within our published protocol (19)). These men were inoculated with wild type *Neisseria gonorrhoeae* strain FA1090, clinically followed daily in an inpatient setting, and treated with antibiotics upon development of urethritis in accordance with our established protocol (15, 19). Urethritis and indication for treatment was defined as visible urethral discharge observed by the study clinician. However participants could request treatment and termination from the study at any time. Monitoring of infection was performed by daily visual inspection, bacterial quantitation of urine (see below), and enumeration of leukocytes in urine sediment via microscopy (see below). Volunteers whose samples were used in this analysis were controls from two different studies – one group was inoculated with wild type FA1090 lot A25 between 2014 and 2015 (NCT04870138) (35), and the other were inoculated with wild type FA1090 lot A26 between 2017 and 2018 (NCT03840811). Both lots were prepared and extensively characterized according to procedures described in a Chemistry Manufacturing and Controls (CMC) document submitted to the Food and Drug Administration (FDA). It is used in human challenge studies through an investigational new drug (IND) authorization by the FDA (IND 15123) (36).

### Trial Data Management

Upon signing consent, participants were issued unique study code number used on all subsequent study data forms, specimens, and case report forms to ensure de-identification of study data. Documentation linking de-identified study codes to patient-identifiable information is encrypted and only accessible by the study principal investigator. For each participant a study documentation form labeled with the individual study code was utilized to record all performed procedures and assessments (e.g. time of inoculation, calculated inoculum administered, bacterial quantitation of urine samples) during the study. Study codes were additionally replaced with arbitrary patient designation numbers (e.g Participant 101, 102…) for publication of this data set. These practices were performed under supervision of the principle investigator with approval and oversight by the University of North Carolina Data and Safety Monitoring Board.

### Specimens

Up to 50mL of first-void urine was obtained from volunteers at three timepoints: 1. Up to 30 days prior to urethral inoculation; 2. On the day of symptomatic urethritis prior to administration of antibiotics; 3. At a follow-up visit 3 to 7 days after treatment to confirm test of cure. Urine was self-collected from patients in specimen cups and immediately given to study personnel for processing. 10mL of urine was centrifuged in a conical tube at room temperature at 400g for 10 minutes, decanted, and the remaining pellet resuspended in residual supernatant to perform pyuria quantitation as previously described (19). The remaining urine (∼40mL) was transferred to a conical tube and spun at 1500g for 10minutes at 4°C within 15 minutes of collection. Urine supernatant was snap frozen on dry ice, and subsequently placed in a -80°C refrigerator, and the remaining pellet was resuspended in gonococcal base (GCB) broth for quantitation of bacteriuria as previously described (19). Plasma was derived from whole blood by venipuncture that was collected in heparin-treated vials. Blood was spun at 1500*g* for 10 minutes at 4°C. Plasma was removed from the whole blood supernatant, snap frozen on dry ice, and later placed in a -80°C refrigerator. Samples were later shipped to Duke University on dry ice, stored at -80°C until use, and thawed on ice once at time of analysis (6-30-2015 and 08-17-2021)

### Cytokine analysis

Cytokines were quantified using Milliplex magnetic Luminex bead arrays (Millipore Sigma). Plasma samples were assayed using a 38-plex kit (# HCYTMAG-60K-PX38) and urine using 41-plex kit (# HCYTMAG-60K-PX41). The 38-plex assayed soluble CD40 Ligand (sCD40L), Epidermal Growth Factor (EGF), Eotaxin (CCL11), Fibroblast Growth Factor-2 (FGF-2), Fms-like Tyrosine Kinase 3 Ligand (Flt-3L), Fractalkine (CX3CL1), Granulocyte Colony-Stimulating Factor (G-CSF), Granulocyte-Macrophage Colony-Stimulating Factor (GM-CSF), Growth-Related Oncogene (detects GROα/β/γ, also known as CXCL1/2/3), Interferon-α2 (IFN-α2), IFN-γ, Interleukin (IL)-1α, IL-1β, IL-1 receptor antagonist (IL-1RA), IL-2, IL-3, IL-4, IL-5, IL-6, IL-7, IL-8/CXCL8, IL-9, IL-10, IL-12p40, IL-12p70, IL-13, IL-15, IL-17A, IFN-γ-induced-protein-10 (IP-10/CXCL10), Monocyte Chemoattractant Protein-1 (MCP-1/CCL2), MCP-3 (CCL7), Macrophage-Derived Chemokine (MDC/CCL22), Macrophage Inflammatory Protein-1α (MIP-1α/CCL3), MIP-1β (CCL4), Transforming Growth Factor-α (TGF-α), Tumor Necrosis Factor-alpha (TNF-α), TNF-β, and Vascular Endothelial Growth Factor (VEGF), while the 41-plex included these 38 plus Platelet-Derived Growth Factor-AA (PDGF-AA), PDGF-AB/BB, and Regulated on Activation Normal T Cell Expressed and Secreted (RANTES / CCL5). Plasma samples were assayed according to the manufacturer’s instructions. Urine samples were neutralized using PIPES buffer (Sigma) to a final concentration of 25mM. PBS was used as a matrix for the assay standards with PIPES+PBS included as a negative control. Assays were then completed according to the manufacturer’s instructions. Assays were read using a Luminex 100 or FlexMAP3D instrument (Luminex corp.) and analyzed using BioPlex Manager (Bio-Rad). Cytokine concentrations were interpolated from standard curves generated using manufacturer-provided standards. Values above the upper limit of quantitation were imputed for analysis as the upper limit of quantitation. Values below the lower level of quantitation (LLOQ; left-censored values) were imputed as half the plate- and analyte-specific LLOQ. As the participants studied were part of two separate clinical trials at two timepoints (2015, 2021), the samples were assayed in two separate batches. Raw and imputed values by assay batch, urine bacterial quantitation and pyuria quantitation are in S1 Table.

### Statistical testing

All statistical testing was performed using GraphPad Prism version 10.5. For differences between timepoints, interpolated cytokine concentrations were compared using a two-tailed Wilcoxon-paired signed rank test with Pratt correction for ties values. This was chosen given the degree of data censorship due to values falling outside the limits of quantitation (up to 70%), which complicates log-normalization that might otherwise allow for parametric testing (37, 38). While these tests substituted left-censored values with half the LLOQ for analysis, re-analyzing the data after substituting these values with zero or LLOQ–0.1 did not substantially affect hypothesis testing (data not shown). We did not perform multiple comparisons testing, as these immune markers are unlikely to be independent of each other, and no formal hypothesis testing was intended for this exploratory study (37, 39). For comparisons in Early and Late group analysis, we used a Mann-Whitney U test. For correlations between urine bacterial counts, urine sediment neutrophil counts, and urine cytokines, values were log_10_ transformed to improve assumptions of a normal distribution, and Pearson correlations were performed.

## Data Availability

The Excel file including all multiplex reads, bacterial counts, and neutrophils counts used for all analyses in this paper is attached as Table S1. Additionally, it will be published to the UNC Dataverse at time of acceptance.

https://doi.org/10.15139/S3/XJUSBZ

## Acknowledgements

This project was supported through funding through the National Institute of Allergy and Infectious Disease (U01-AI114378, U19AI031496) and the National Center for Advancing Translational Sciences (KL12-TR004416). Cytokine profiling was performed in the Regional Biocontainment Laboratory at Duke University, which received partial support for construction and renovation from the National Institutes of Health (UC6-AI058607 and G20-AI167200), and facility support from the National Institutes of Health (UC7-AI180254). We thank Ryan Wuerker and Melissa Samo for technical assistance.

## Supporting Information

**S1 Fig. Graphic representation of urine cytokines with >70% values below the lower limit of quantitation (LLOQ).**

Each dot represents a cytokine measurement from one participant, with lines connecting measurements from the same participant. Values below the LLOQ (dotted line) are displayed as one half LLOQ and unresolved from other non-detects.

**S2 Fig. Additional Comparisons in Acute Infection Urine Cytokine Levels in Early and Late Presenters.**

Each dot represents a cytokine measurement from one participant. Values below the lower limit of quantitation (LLOQ, dotted line) are displayed as one half LLOQ. Comparison bars indicate statistical significance at *p<0.05; via Mann-Whitney U Test.

**S3 Fig. Additional linear correlation plots between Acute Infection timepoint urine measurements of all participants.**

All values are log transformed. Symbols represent individual patients. Individual values below the lower limit of quantitation are represented with an “x” rather than a dot. Slopes were not constrained to include the origin.

**S4 Fig. Graphic Representation of serum cytokines with >50% values above the lower limit of quantitation (LLOQ).**

Each dot represents a cytokine measurement from one participant, with lines connecting measurements from the same participant. Values below the LLOQ (dotted line) are displayed as one half LLOQ and unresolved from other non-detects.

**S1 Table. Milliplex Raw Data.**

Tab 1 provides cytokine analyte annotation. Tab 2 provides analyte reads per plate, followed by combined data organized by time point. Limits of detection, and Log transformations, Metadata, and urine CFU, PMN counts are also provided. Red Values denote values outside limits of quantitation, with those values below the limit of quantitation expressed as half the limit of detection for that plate and analyte.

## Author Contributions

MPM performed Formal Analysis, Visualization, and Writing the Original Draft, Reviewing and Editing. MMH performed Conceptualization, Data Curation, Formal Analysis, Investigation, Reviewing and Editing. AW performed Supervision, Reviewing and Editing. ANM performed Data Curation, Investigation, Supervision, Validation, Reviewing and Editing. JAD performed Conceptualization, Funding Acquisition, Investigation, Methodology, Project Administration, Supervision, Reviewing and Editing.

